# The Impact of COVID-19 Restrictions on Childhood Vaccination Uptake: A Rapid Review

**DOI:** 10.1101/2021.06.25.21259371

**Authors:** C Heneghan, J Brassey, T. Jefferson

**Author notes:** https://www.tripdatabase.com. **COLLATERAL global**.

## Abstract

**Background:** Vaccines are highly effective for preventing a range of childhood infections. However, there have been concerns about an alarming decline in vaccinations in 2020 due to the COVID-19 pandemic.

**Methods:** We performed a rapid review for studies that assessed childhood vaccination uptake during restrictive phases of the COVID-19 pandemic.

**Results:** We found 35 published studies that compared changes in the pattern of childhood vaccinations before and during the pandemic. Thirteen were surveys; two mixed-methods surveys and interviews, three modelling studies and 17 retrospective cohort studies with historical controls. We also included ten reports by national or international agencies that had original data on vaccination uptake. Significant global disruptions to vaccine services were reported in Africa, Asia, America (including Latin America and the Caribbean) and Europe. We also found evidence of significant disruption to vaccine uptake for diphtheria tetanus pertussis, BCG, measles and polio. Countries where vaccination rates were already suboptimal had greater drops in uptake and there was evidence of smaller declines in younger children compared to older children. Children born to women who could not read and write were more likely to be incompletely immunized. Various initiatives were used to drive up vaccination rates post restrictions.

**Conclusions:** Obstacles to the delivery of vaccination services during the Covid-19 pandemic drove down immunisation rates, especially in disadvantaged people and poorer countries.

## Introduction

Vaccines have proven to be highly effective at preventing disease and deaths associated with a range of childhood infections. Many childhood diseases that used to be common, including diphtheria, measles, mumps, rubella, pertussis, polio and tetanus, can be prevented by vaccination. The distribution and uptake of vaccines has improved across the globe. In 2019, 86% of infants worldwide received three doses of diphtheria, tetanus, pertussis (DTP3) vaccines, and childhood immunisation is estimated to prevent 2 to 3 million annual deaths from infectious diseases. ^**[1]**^

Despite their effectiveness, the WHO and UNICEF warned of an ‘alarming decline’ in vaccinations in 2020 due to the COVID-19 pandemic. Data from the first four months highlighted a substantial drop in the number of children completing three doses of the DTP3 vaccine - the first time in 28 years global reductions were seen. ^**[2]**^

UNICEF regards protection from vaccine-preventable diseases as a child’s fundamental right and states that right now is ‘time to vigorously monitor the impact on immunisation and plan for services that reach the most vulnerable once restrictions are lifted.’^**[3]**^ We, therefore, set out to synthesise the published literature on the impact of the COVID-19 pandemic restrictions on vaccination coverage for children.

## Methods

We performed a rapid review using a flexible framework for restricted systematic reviews. ^**[4]**^ For the initial search, we restricted the results to peer-reviewed articles using the LitCovid database. LitCovid is a curated literature hub for tracking up-to-date scientific information about SARS-CoV-2 (https://www.ncbi.nlm.nih.gov/research/coronavirus/). It is a comprehensive resource on the subject, providing central access to relevant articles in PubMed. We searched for various terms/phrases associated with childhood vaccinations, e.g., ‘vaccines’, ‘vaccination’ or ‘immunisation’ and ‘children’ or ‘infants’. We screened the title and abstract for inclusion and extracted data into templates on the study identifier, country, the type of vaccination, the study type, the age of the included populations, the primary methods and the main results. We did not formally assess quality as studies were retrospective reviews of records that involved an active lockdown phase and a historical control period, surveys or modelling studies. The quality of included studies was instead assessed using an adapted version of the Oxford Centre for Evidence-Based Medicine Levels of Evidence (https://www.cebm.ox.ac.uk/resources/levels-of-evidence/ocebm-levels-of-evidence)

We also included reports by national or international agencies that included original data on vaccination uptake. We summarised data narratively and reported the outcomes as stated, including quantitative estimates, where feasible and relevant. We presented the data by disruption of services and by diphtheria tetanus pertussis, BCG, measles and polio vaccine uptake. We also extracted up to date numbers from the Global Polio Eradication Initiative (GPEI) database to assess the latest impact of polio vaccination disruptions in Afghanistan and Pakistan, where polio is endemic (https://polioeradication.org/). Our review process, strategy and rationale can be accessed at https://collateralglobal.org/article/what-is-a-rapid-review/.

## Results

Figure 1 reports the Identification of Childhood Vaccination studies via databases. From 389 records identified, we found 35 published studies that compared changes in the pattern of childhood vaccinations before and during the pandemic. 32 studies assessed evidence from 22 countries: Afghanistan, Bangladesh, China, Germany, India, Indonesia, Japan, Kuwait, The Netherlands, Portugal, Rwanda, Scotland, Senegal, Sierra Leone, South Korea, Spain (16 countries, one study each); England, Ethiopia, Italy, Pakistan, Saudi Arabia (five countries, two studies each). For the USA, there were six studies. Three studies reported on multiple countries: two were done in Africa [Masresha 2020 and Abbas K 2020] and one in South-East Asia and the Western Pacific (SEAR/WPR) [Harris 2021].

**Figure 1.**
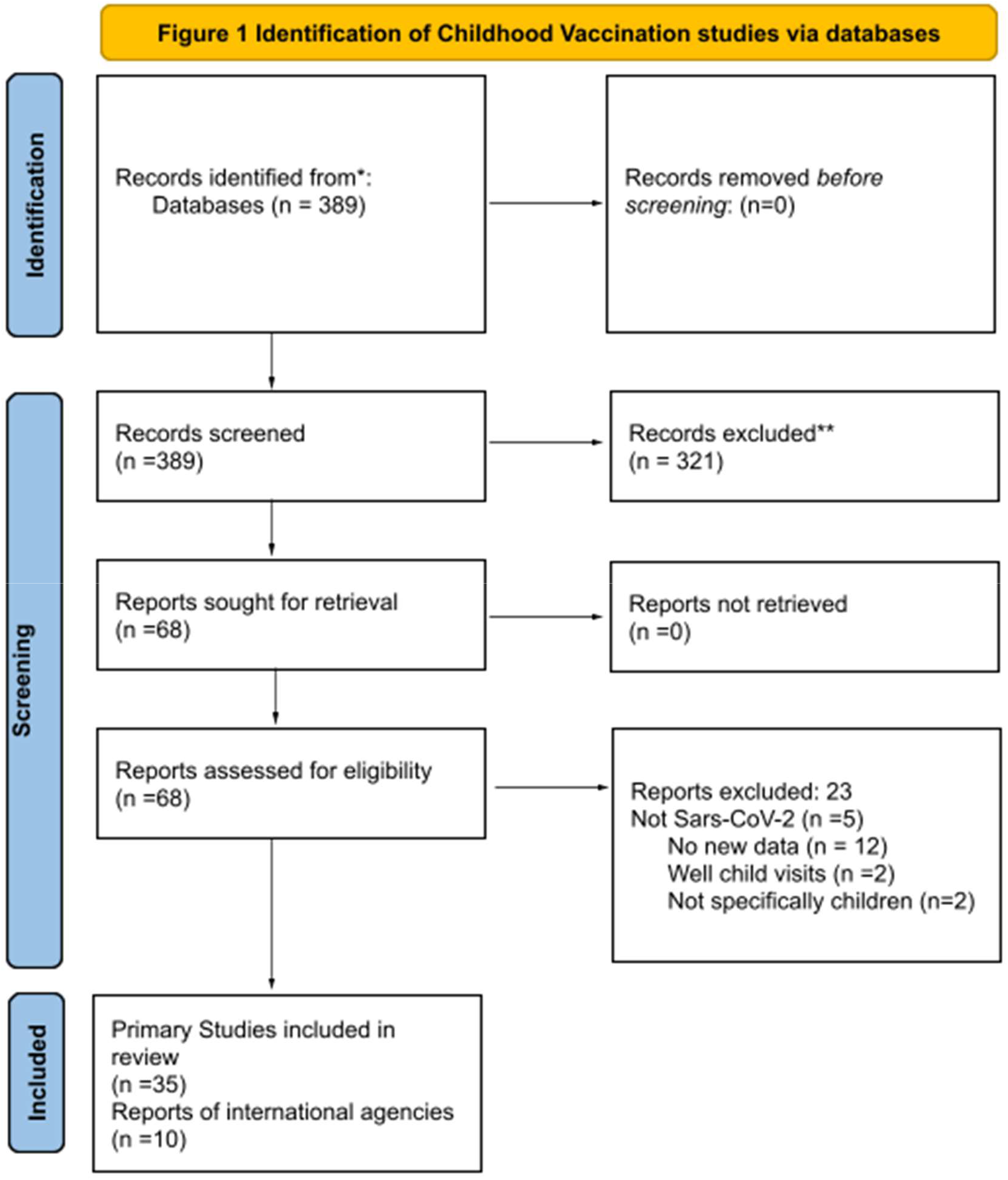
PRISMA 2020 Flow Diagram. *Consider, if feasible to do so, reporting the number of records identified from each database or register searched (rather than the total number across all databases/registers). **If automation tools were used, indicate how many records were excluded by a human and how many were excluded by automation tools. *From:* Page MJ, McKenzie JE, Bossuyt PM, Boutron I, Hoffmann TC, Mulrow CD, et al. The PRISMA 2020 statement: an updated guideline for reporting systematic reviews. BMJ 2021;372:n71. doi: 10.1136/bmj.n71 For more information, visit: http://www.prisma-statement.org/

**Figure 2.**
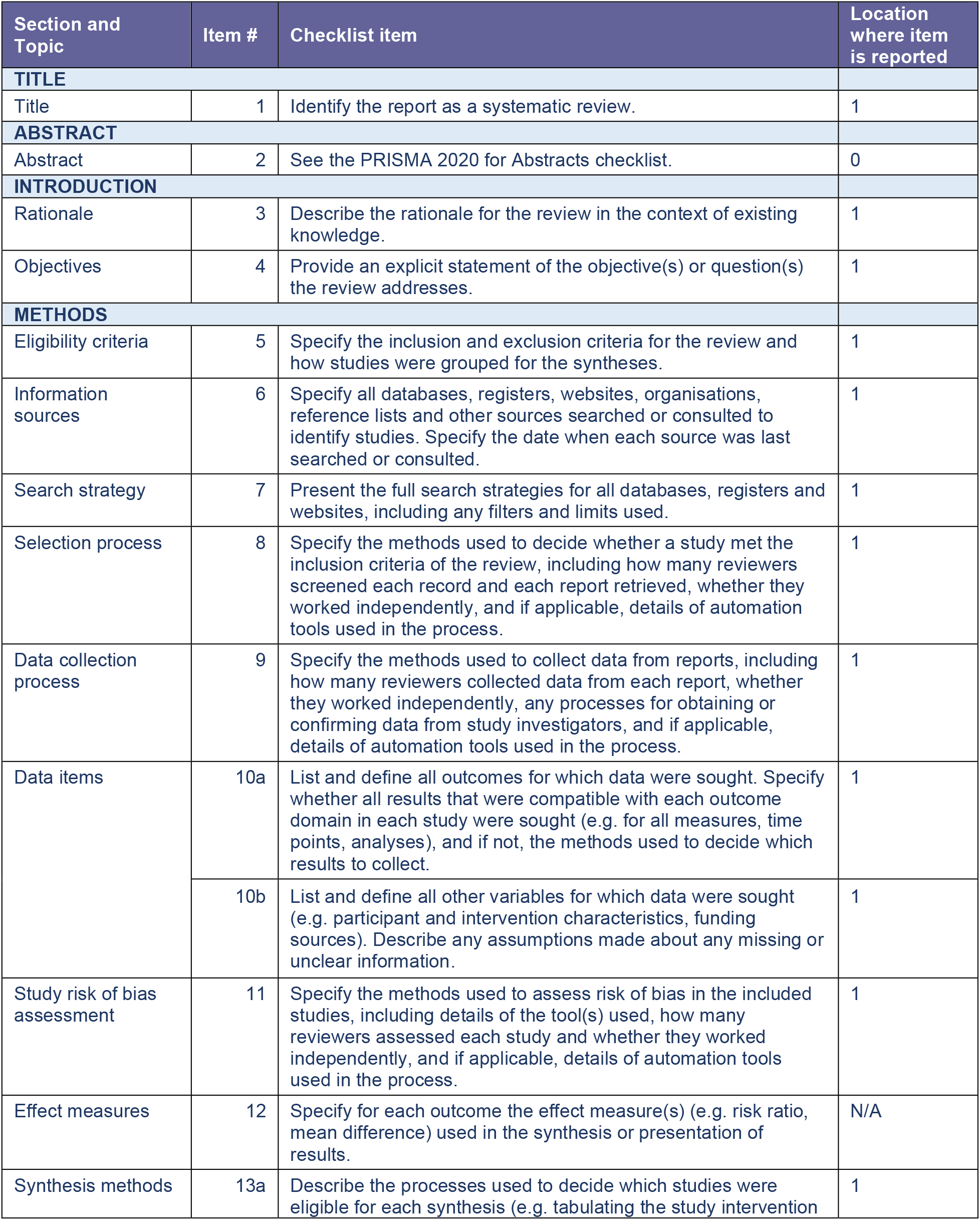
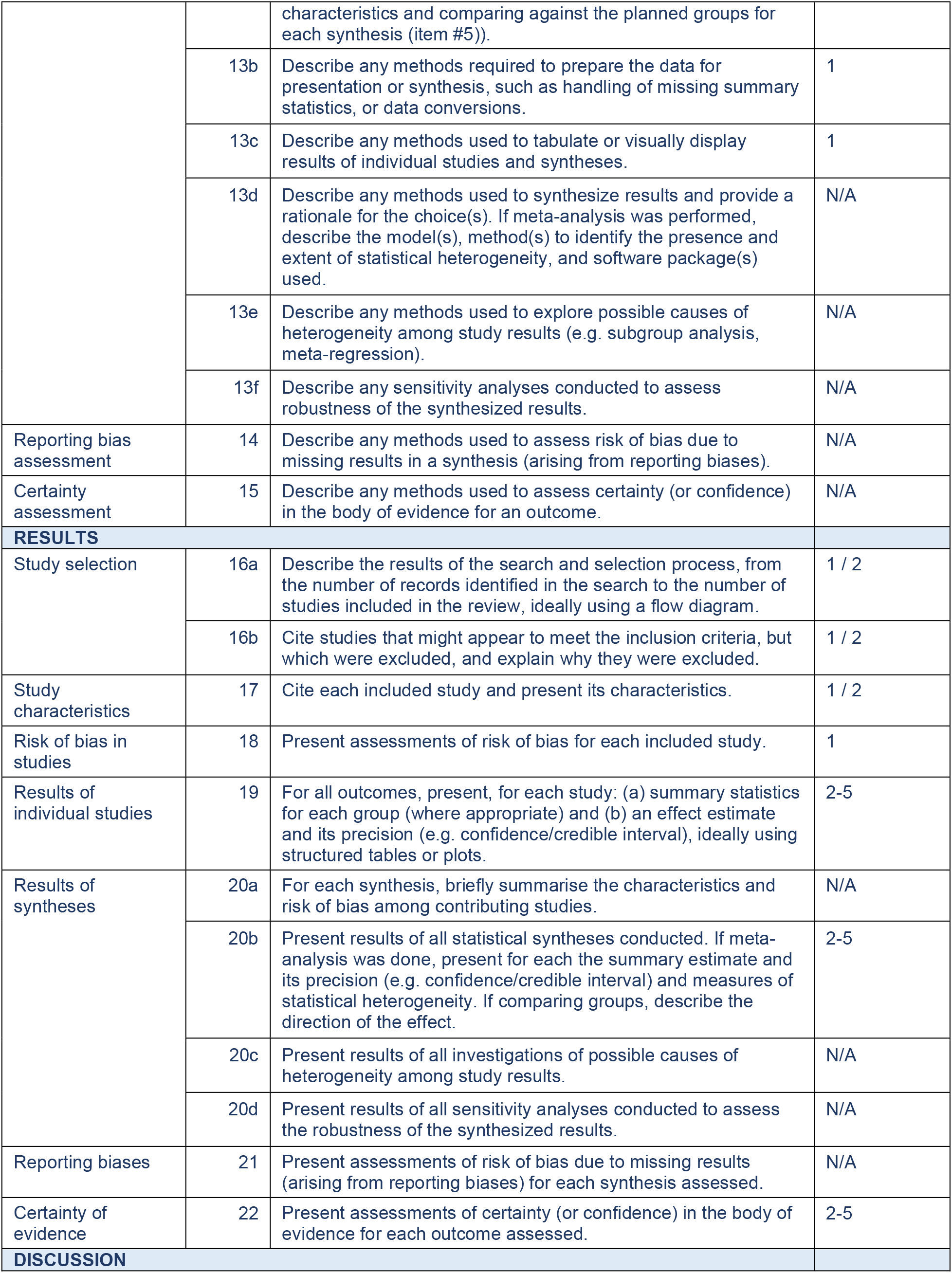
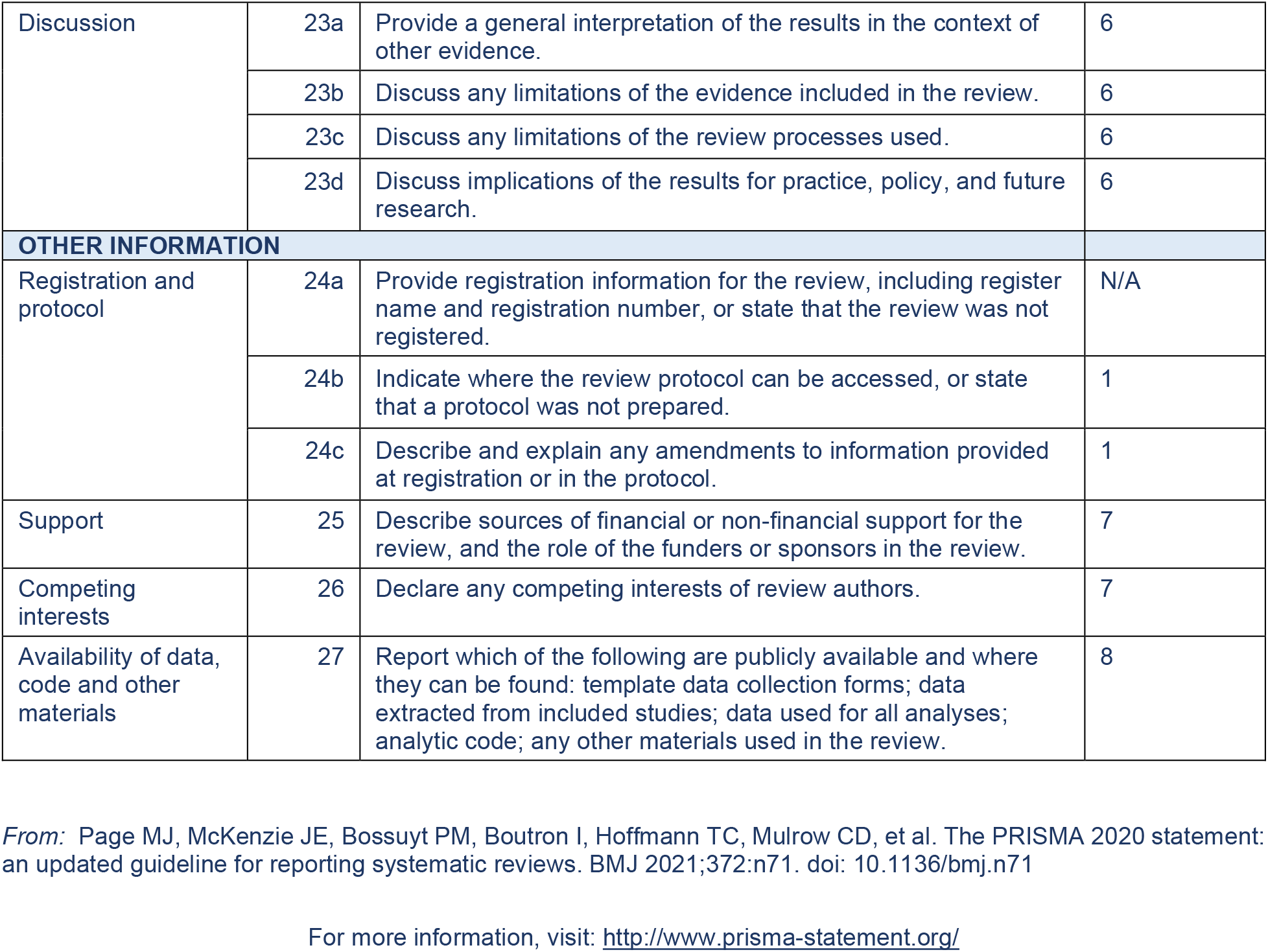
PRISMA Checklist.

In terms of methods, thirteen studies were surveys; two were mixed-methods surveys and interviews, three studies were models (one provided data on a retrospective cohort), and 17 were retrospective cohort studies.

We also included ten reports by national or international agencies that had original data on vaccination uptake: one each for Blue Cross, the Nuffield Trust, PAHO and Public Health England, two reports for UNICEF (one jointly with PAHO) and four WHO reports. (See TABLE 2)

**TABLE 1:**
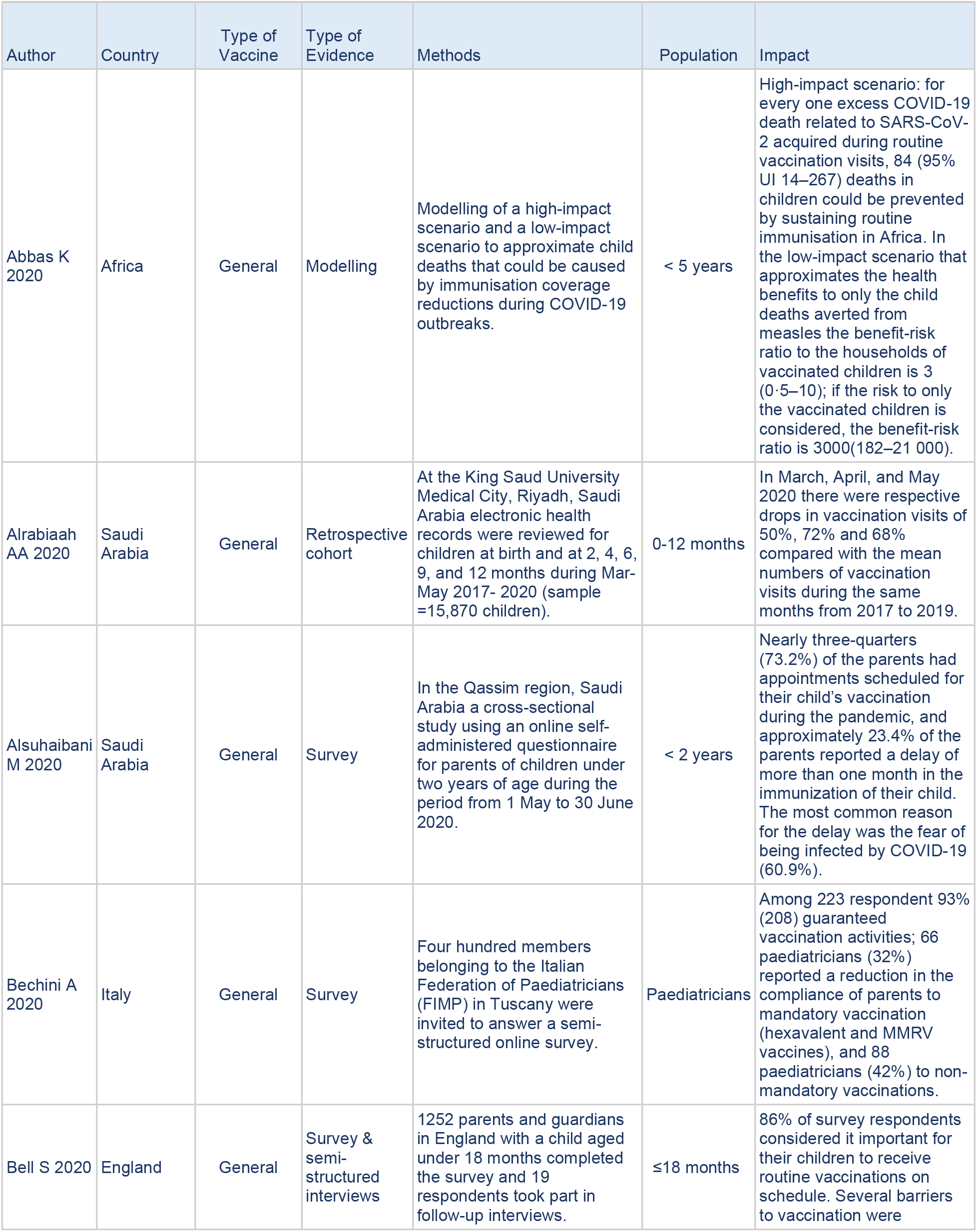
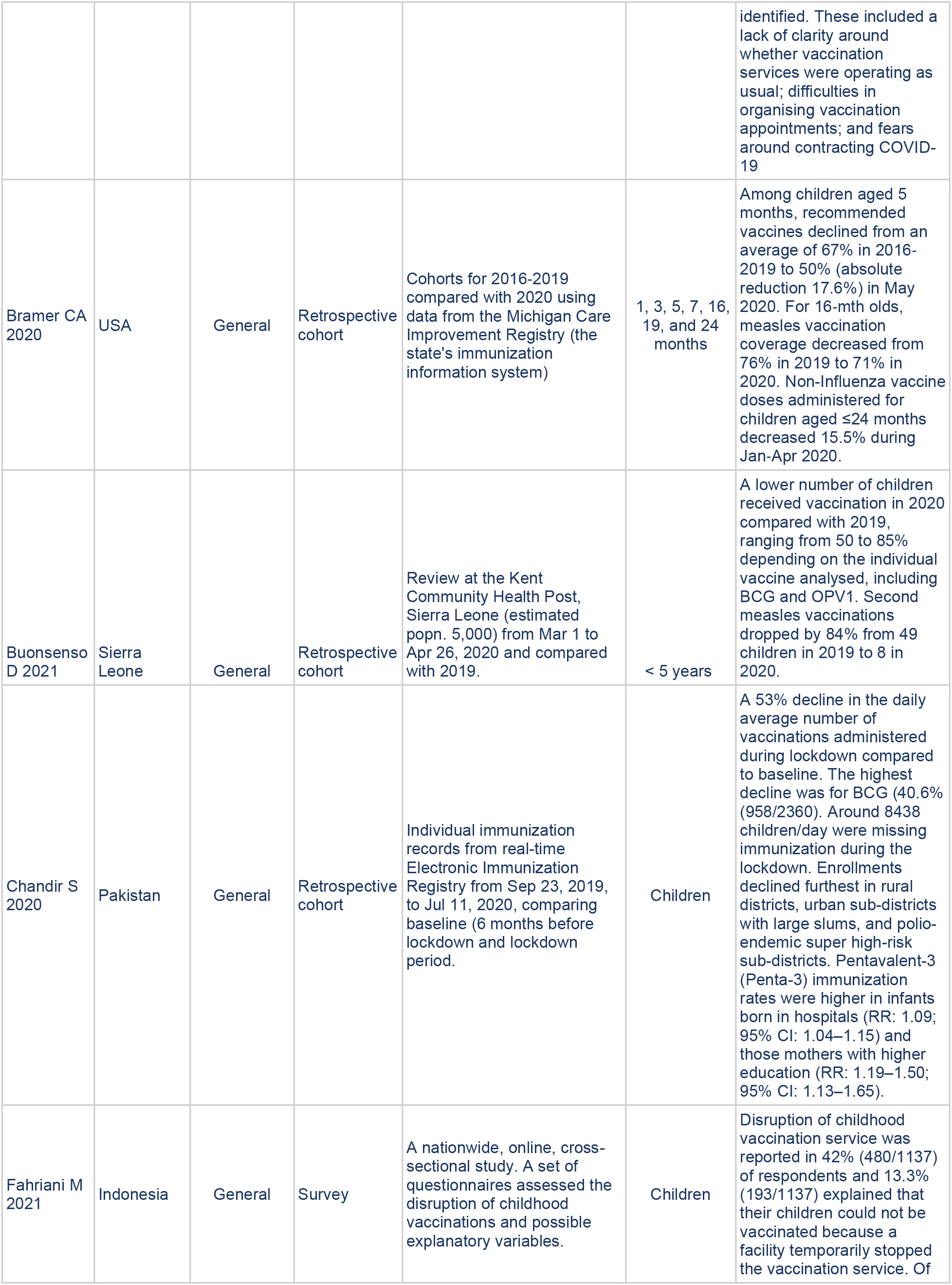
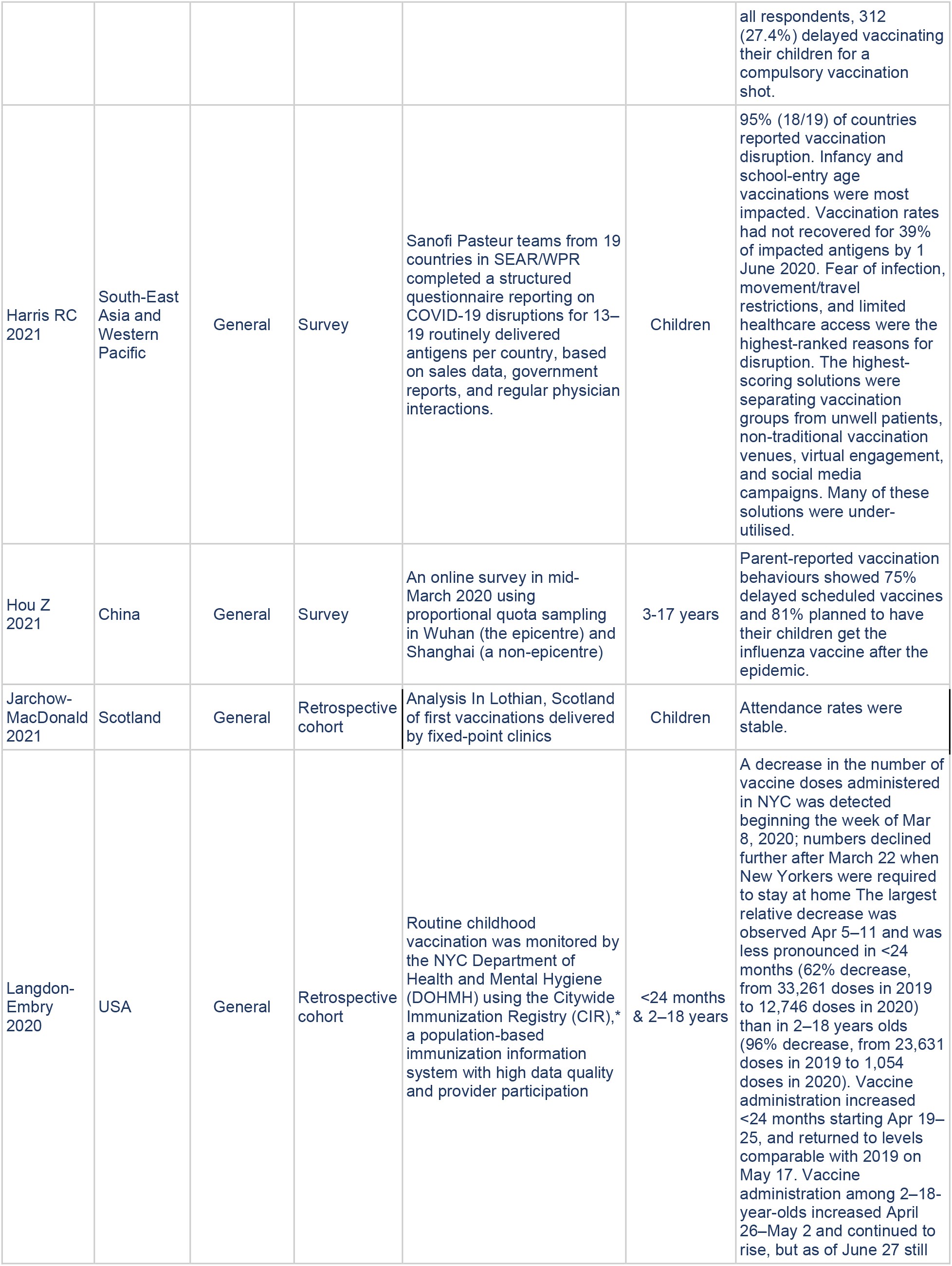
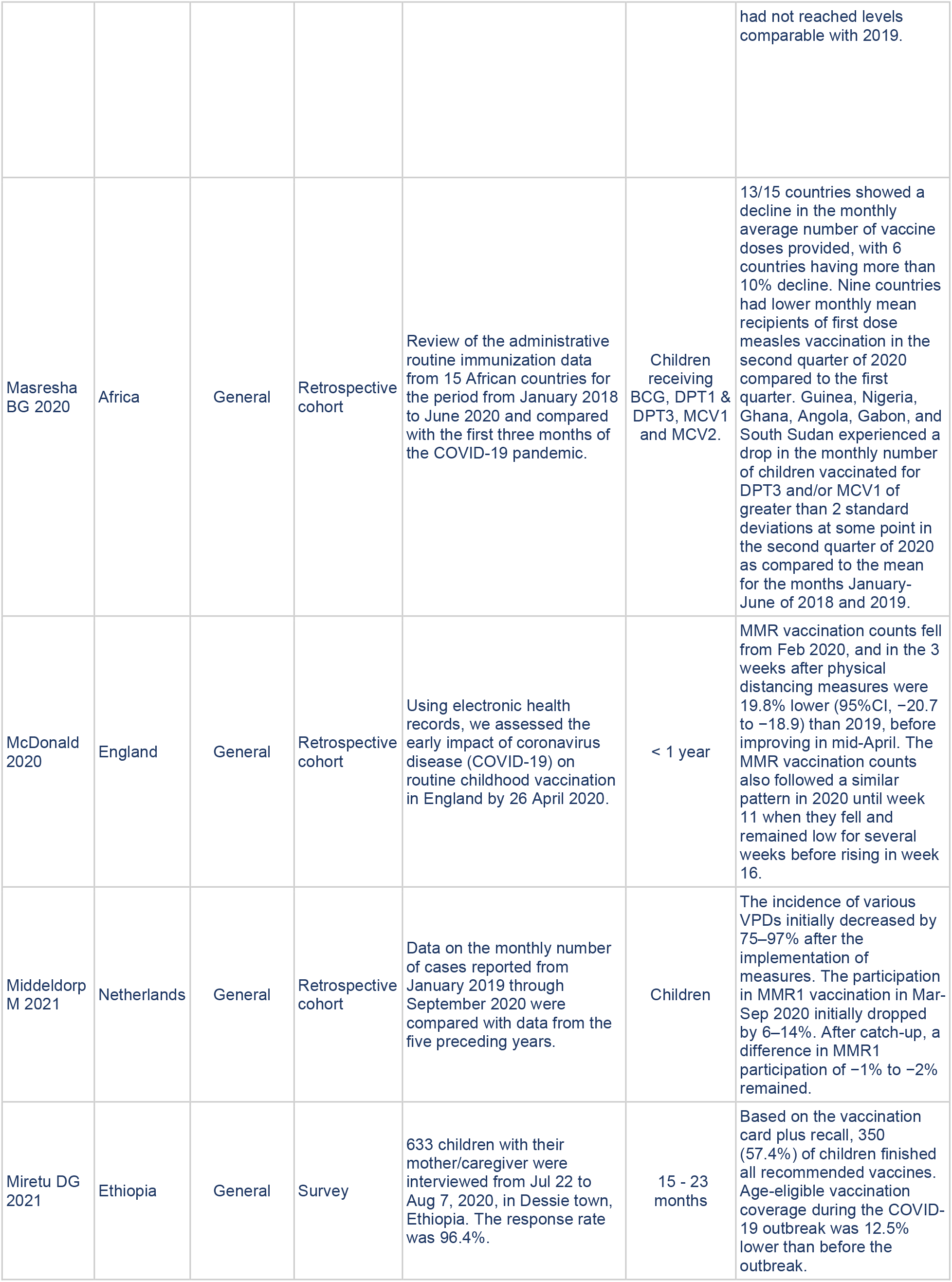
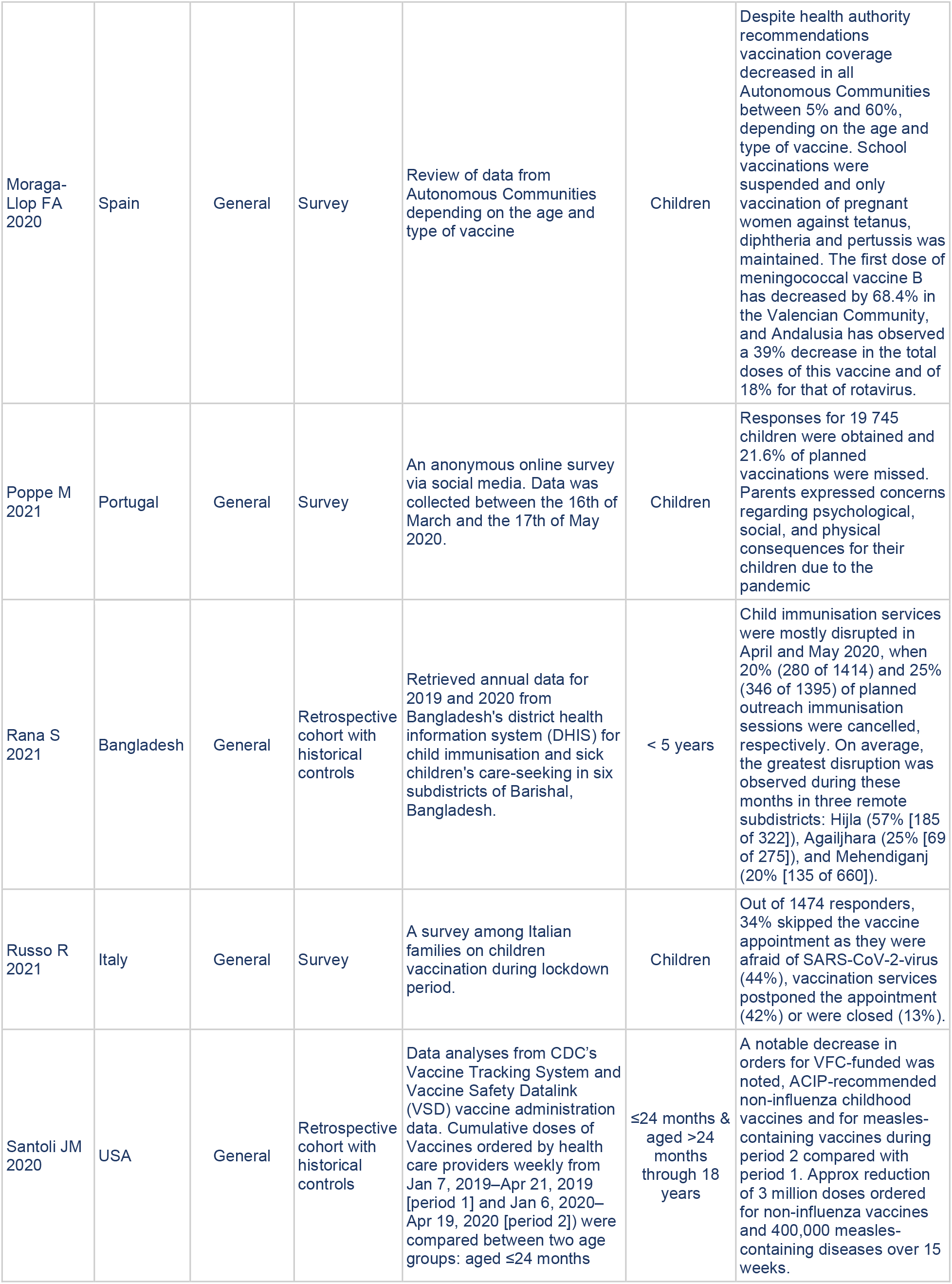
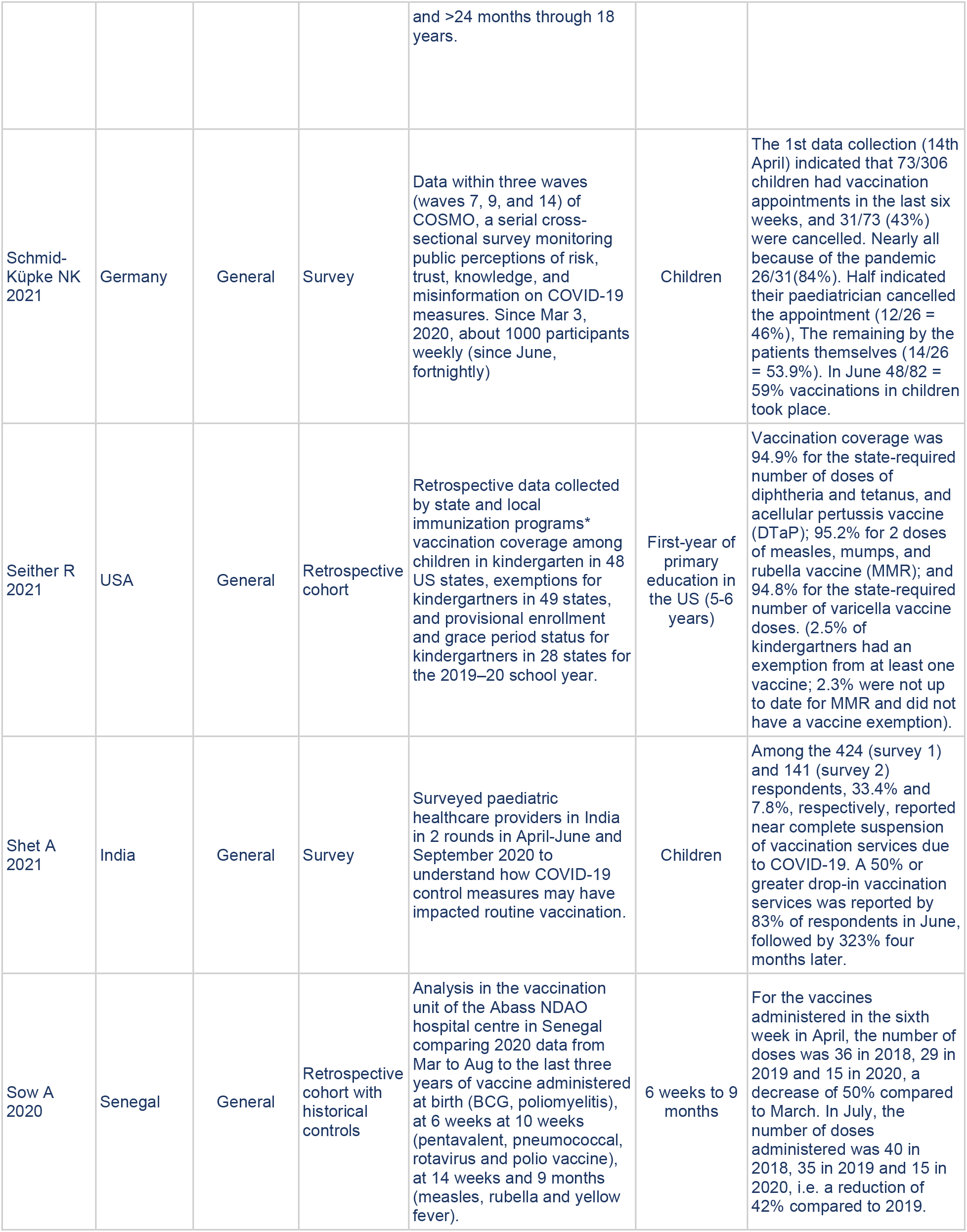
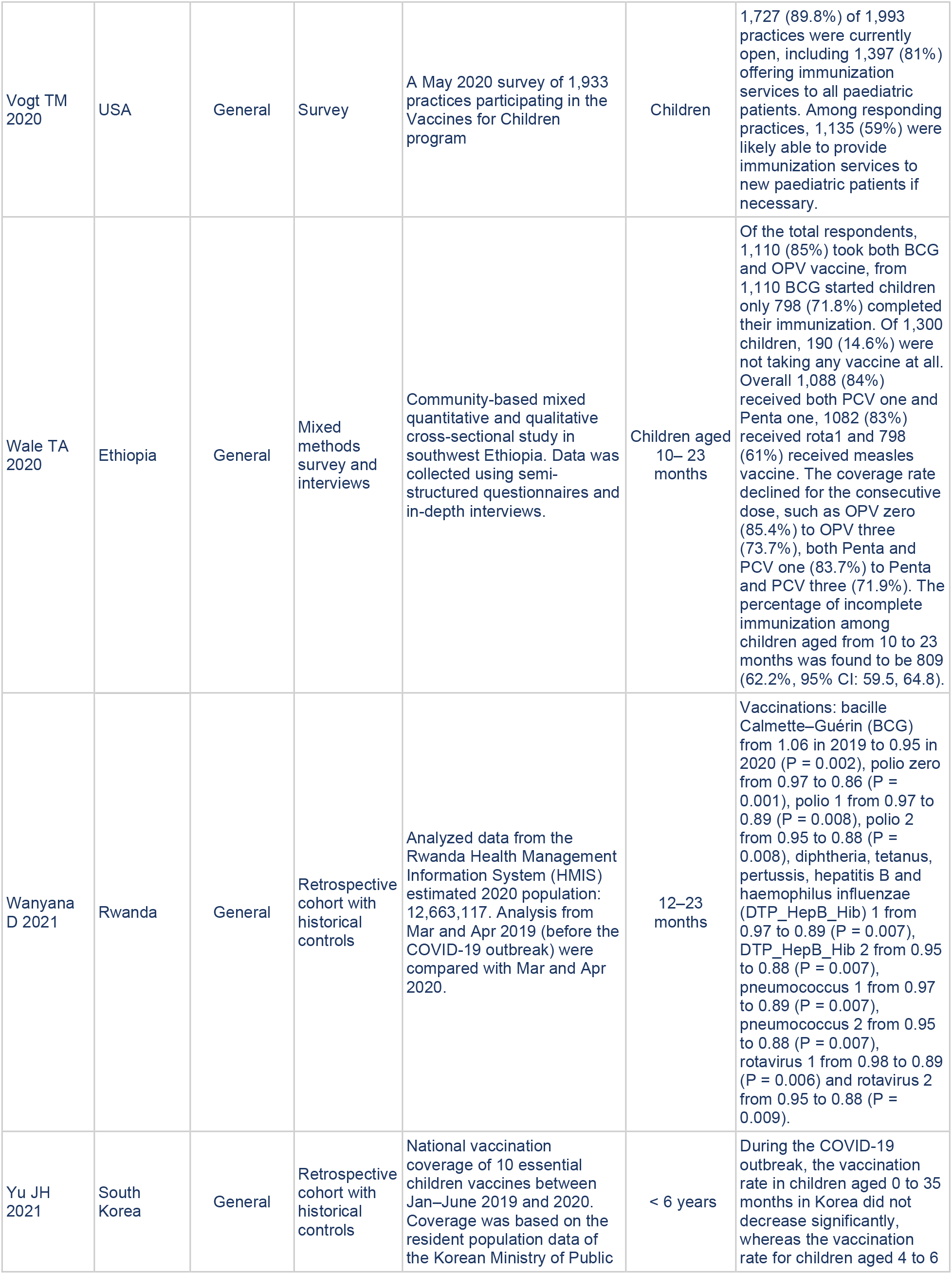
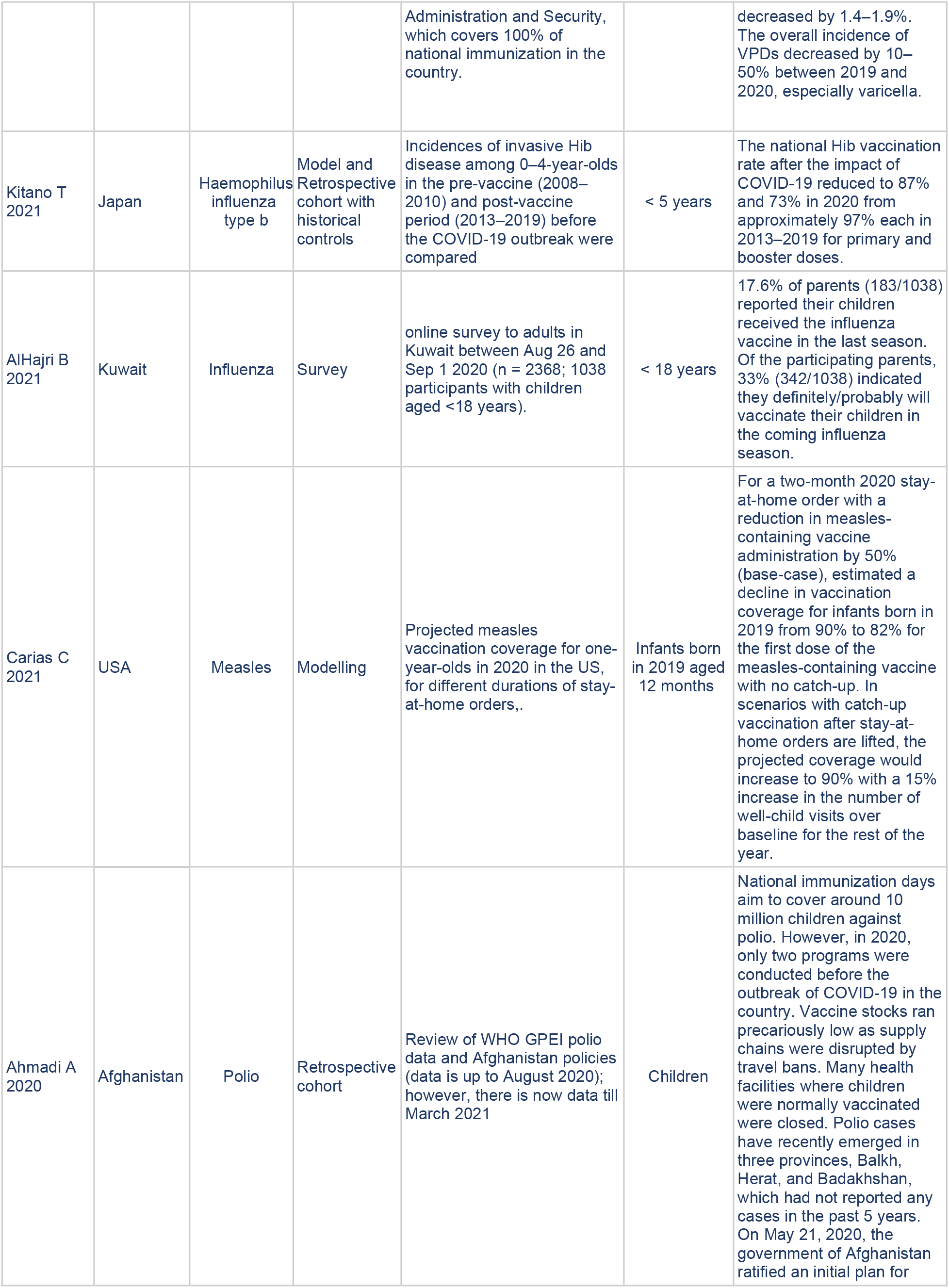
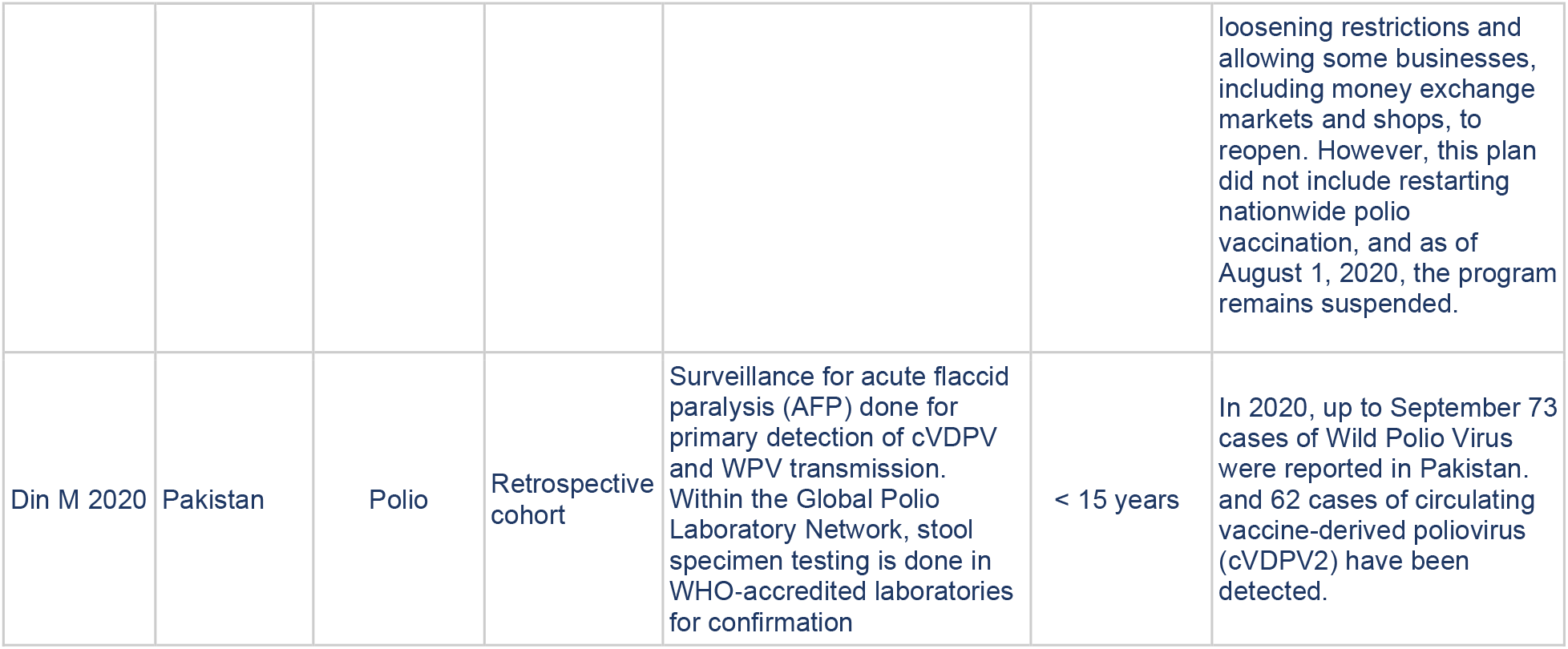
Primary Studies on Childhood Vaccination Uptake During the COVID-19 Pandemic.

**TABLE 2:** Reports by National and International Agencies on Childhood Vaccination Uptake During the COVID-19 Pandemic.

| Organization and Year of Publication | Title | Summary | Link |
| --- | --- | --- | --- |
| Blue Cross 2020 | Blue Cross Blue Shield. Missing vaccinations during COVID-19 puts our children and communities at risk | Reports up to 26% drop in MMR, DTP & polio vaccines between Jan-Sept 2020 in America. | https://www.bcbs.com/the-health-of-america/infographics/missing-vaccinations-during-covid-19-puts-our-children-and-communities-at-risk |
| Nuffield Trust 2021 | Vaccination coverage for children and mothers | Indicators that look at vaccination coverage for children and mothers in the UK and internationally during the coronavirus (Covid-19) pandemic, coverage was largely maintained when compared to coverage in 2019/20. | https://www.nuffieldtrust.org.uk/resource/vaccination-coverage-for-children-and-mothers-1 |
| Public Health England 2021 | Impact of COVID-19 on childhood vaccination counts to week 4 in 2021, and vaccine coverage to December 2020 in England: interim analyses | Hexavalent and MMR vaccination counts fell at the time of the introduction of physical distancing measures in March 2020 (week 13) compared to the same period in 2019. This was followed by a rise from weeks 16 onwards which has stabilised and is comparable to vaccination counts prior to the COVID-19 pandemic. | Impact of COVID-19 on childhood vaccination counts to week 4 in 2021, and vaccine coverage to December 2020 in England: interim analyses |
| UNICEF 2020 | Challenges posed by the COVID-19 pandemic in the health of women, children, and adolescents in Latin America and the Caribbean | During the pandemic, the population's fear of COVID-19 and public transport limitations and confinement and physical distance policies led to a decrease in the demand for vaccination services in half of the 38 countries in the region that report information to the Pan American Health Organization (PAHO). | https://www.unicef.org/lac/media/16376/file/undp-rblac-CD19-PDS-Number19-UNICEF-Salud-EN.pdf |
| PAHO* 2020 | Summary of the Status of National Immunization Programs during the COVID-19 Pandemic, July 2020. |  | https://iris.paho.org/handle/10665.2/52544 |
| UNICEF 2020 | Immunization coverage Are we losing ground? | Lockdown measures had substantially hindered the delivery of immunization services in at least 68 countries, putting approximately 80 million children under the age of 1 at increased risk of contracting vaccine-preventable diseases. | https://data.unicef.org/resources/immunization-coverage-are-we-losing-ground/ |
| WHO pulse interim survey 2020 | Pulse survey on continuity of essential health services during the COVID-19 pandemic: interim report, 27 August 2020 | 105 countries responded. Severe/complete disruption of outreach routine mobile immunization services occurred in 18% of 91 countries and in 10% disruption to static routine immunization occurred. | https://www.who.int/publications/i/item/WHO-2019-nCoV-EHS_continuity-survey-2020.1 |
| WHO pulse second-round survey 2021 | The second round of the national pulse survey on continuity of essential health services during the COVID-19 pandemic | In January-March 2021 for 135 countries surveyed more than one-third were reporting disruptions to immunization services. | https://www.who.int/publications/i/item/WHO-2019-nCoV-EHS-continuity-survey-2021.1 |
| WHO pulse survey 15 May report 2020 | At least 80 million children under one at risk of diseases such as diphtheria, measles and polio as COVID-19 disrupts routine vaccination efforts, warn Gavi, WHO and UNICEF | More than half (53%) of the 129 countries where data were available reported moderate-to-severe disruptions or a total suspension of vaccination services during March-April 2020. | https://www.who.int/news/item/22-05-2020-at-least-80-million-children-under-one-at-risk-of-diseases-such-as-diphtheria-measles-and-polio-as-covid-19-disrupts-routine-vaccination-efforts-warn-gavi-who-and-unicef |
| WHO Niger 2020 | Niger reports a new polio outbreak. | Niger has reported a new polio outbreak that has affected two children in Niamey and Tillaberi region, | https://www.afro.who.int/news/niger-reports-new-polio-outbreak. |

\* Pan American Health Organization

### Disruption to services

Data collated by the WHO, UNICEF, Gavi, and the Sabin Vaccine Institute show that pandemic restrictions substantially reduced the delivery of immunization services in at least 68 countries affecting over 80 million children under the age of one. [UNICEF 2020] According to the WHO’s first pulse interim survey published in August 2020, 16/91 (18%) of countries reported severe/complete disruption of routine mobile immunization services, and 10% reported disruption to static routine immunisation services. About half of the countries reported partial disruptions of routine immunisation for both health facilities and mobile services. [WHO first-round survey 2020] On May 15, the pulse survey reported that more than half (53%) of the 129 countries with available data had moderate-to-severe disruptions or a total suspension of vaccination services from March to April 2020. [WHO pulse survey 15 May 2020] The WHO’s second round national pulse survey from January to March 2021 reported that more than one-third of 135 countries experienced disruptions to immunisation services: routine facility-based disruption occurred in 35 (34%) countries surveyed and outreach immunisation services occurred in 35 (39%) countries. [WHO second-round survey 2021]

### Africa

A review of the administrative routine immunisation data from 15 **African** countries in 2020 revealed that 13 countries experienced declines in the monthly average number of vaccine doses provided from January to June compared with 2018 and 2019. [Masresha BG 2020]. In **Ethiopia**, 633 children and their mother/caregiver were interviewed from Jul 22 to Aug 7, 2020. Based on their recall, plus vaccination cards, 350 (57%) children finished all recommended vaccines. Vaccination coverage during the outbreak was 12.5% lower than before the outbreak. [Miretu DG 2021]

### Asia

Twenty percent of child immunisation services in **Bangladesh** were cancelled in April 2020 and 25% in May. The most significant disruption occurred in remote subdistricts: Hijla (57%), Agailjhara (25%) and Mehendiganj (20%). Improved coverage appeared in the post-disruption months (July to October 2020), with about 99% of immunisation sessions held. [Rana S 2021] In **India**, survey data of paediatric healthcare providers from April to June and September 2020 reported a greater than 50% drop in vaccination services by 83% of the respondents in June (n=424 respondents) and 33% in September (n=141). [Shet 2021] In **Indonesia**, 42% (480/1137) of survey respondents reported disruption of childhood vaccination services in local health facilities and 13% (193/1137) of respondents explained that their children could not be vaccinated because a healthcare facility temporarily stopped vaccination service. Of all respondents, 312 (27%) delayed vaccinating their children for a compulsory vaccination shot. [Fahriani M 2021] In **South Korea**, during the outbreak, the vaccination rate in children aged <35 months did not decrease significantly, whereas the vaccination rate for children aged 4 to 6 decreased by 1.4 to 1.9%. [Yu 2020] In **Japan**, COVID-19 resulted in decreases in the vaccination rate for Haemophilus influenza type b in <5-year-olds from approximately 97% each in 2013 and 2019 to 87% and 73% in 2020 for primary and booster doses, respectively. [Kitano 2021]

### Middle East

At the King Saud University Medical City, Riyadh, in **Saudi Arabia**, electronic health records reported drops in vaccination visits of 50%, 72% and 68% in March, April, and May 2020 compared with the same months in 2017 to 2019. [Alrabiaah 2020] Furthermore, in the Qassim region of **Saudi Arabia**, a questionnaire for parents of children under two years of age conducted between 1 May to 30 June 2020 reported that 23% had a delay of more than one month in the immunisation of their child. The most common reason for delay was fear of COVID-19 infection. [Alsuhaibani M 2020]

### Europe

Weekly survey data from roughly 1,000 participants in **Germany** reported on 14 April 2020 that 31 of 73 scheduled childhood vaccinations were cancelled within the previous six weeks - 26 (84%) because of the pandemic. Nearly half of the parents (46%) indicated that their paediatrician cancelled the appointment. According to the survey, two-thirds of the cancelled vaccination appointments had been caught up, but appointments had not been scheduled for roughly one in five. [Schmid-Küpke 2021] In Lothian, **Scotland**, fixed-point clinics delivered infants’ first vaccinations at various locations accessible by public transport, leading to stable attendance rates. [Jarchow-MacDonald 2021] In **Spain**, a review of data from Autonomous Communities reported that vaccination coverage in all communities decreased by between 5% and 60%, depending on the age and type of vaccine. The first dose of meningococcal vaccine B decreased by 68.4% in the Valencian Community, and Andalusia observed a 39% decrease in the total doses of this vaccine [Moraga-Llop FA 2020]. In **Portugal**, survey responses for 19,745 children reported that 21.6% of planned vaccinations were missed. [Poppe M 2021] Similarly, a survey among **Italian** families on childhood vaccinations during the lockdown period reported that out of 1474 responders, 34% skipped vaccine appointments because they were afraid of the SARS-CoV-2 virus, the vaccination provider postponed the appointment, or the service was closed. [Russo R 2021]

### USA

**US** CDC Data indicated that three million fewer doses of non-influenza vaccines were ordered from 6 January to 19 April 2020, compared with the previous year. [Santoli JM 2020] A decrease was seen in the number of vaccine doses administered in **New York City** in March and April 2020. In children <24 months of age, the largest relative decrease occurred between 5 to 11 April with a 62% decrease - from 33,261 doses in 2019 to 12,746 doses in 2020. In those aged 2 to 18 years, a 96% decrease was observed - from 23,631 doses in 2019 to 1,054 doses in 2020. Vaccine administration in children <24 months returned to levels comparable with 2019 in May 2020, but in those aged 2 to 18 years, levels had not returned to normal by the end of June. [Langdon-Embry 2020] Blue Cross Blue Shield, which insures 1 in 3 **Americans**, reported a drop of up to 26% in MMR, DTP & polio vaccines between January and Sept 2020. [Blue Cross 2020]

### Latin America and the Caribbean

In **Latin America and the Caribbean**, difficulties caused by public transport limitations, confinement and physical distance policies, along with fear of COVID, led to a decrease in demand for vaccination services in half of the 38 countries that reported information to the Pan American Health Organization [PAHO] in June 2020. ^**[5]**^ [PAHO 2020] Initiatives to increase vaccination rates included institutional drive-through vaccination, mobile vaccination centres, vaccination in homes and strategic locations, follow-up vaccinations using the electronic immunisation registry and emphasis on the importance of maintaining immunisation during a pandemic.

### BCG vaccination

Data from the **Rwanda** Health Management Information System from March and April 2019 showed significant reductions in bacille Calmette– Guérin (BCG) compared with 2020. [Wanyana 2021] In **Pakistan**, there was a 53% decline in the daily average total vaccinations administered. Provincial electronic immunisation registry data reported about 8,400 children per day missed immunisation during the lockdown phase [Chandir S 2020], with the highest decline seen for BCG immunisation 958/2360). **Ethiopia** reported that of 1,110 children who started their BCG vaccination, only 798 (72%) completed their immunisations. [Wale 2020] From January to June 2020, **Burundi, CAR, Chad, DR Congo, Eritrea, Rwanda** and **Senegal** managed to maintain cumulative numbers of vaccinated children for BCG, compared to the mean of 2018/2019. [Masresha BG 2020]

### Diphtheria, Tetanus, Pertussis [DTP]

The **Rwanda** data also showed significant reductions in DTP vaccination rates. [Wanyana 2021] Analysis of children receiving the first and third doses of DTP-containing vaccines in Guinea, Nigeria, Ghana, Angola, Gabon, and South Sudan showed drops in monthly vaccination in the second quarter of 2020 for the third dose compared with 2018-19. **Burundi, CAR, Chad, DR Congo, Eritrea, Rwanda** and **Senegal** managed to maintain the number of vaccinated children for DPT1 and DPT3. [Masresha BG 2020] Declines were seen in Pakistan, with the likelihood of Penta-3 immunisation, which includes the DTP-containing vaccines, reduced by 5% for each week of delay. Retrospective data from a vaccination unit of a hospital centre in **Senegal** comparing data for 2018, 2019 and 2020, reported pentavalent vaccine reductions in ten-week-olds. [Sow A 2020]

### Measles

Eleven studies assessed measles vaccination rates: Abbas 2020, Bechini 2020, Bramer 2020, Carias 2021, Masresha 2020, McDonald 2020, Middeldorp 2021, Santoli 2020, Seither 2021, Sow 2020, and Wale 2020.

Two surveys [Bechini 2020 and Wale 2020] reported reductions in vaccine uptake. Nearly a third (32%) of 223 paediatricians in Tuscany, **Italy**, reported declines in mandatory vaccination, including the measles and rubella vaccine (MMR). Postponements were mainly due to safety fears. [Bechini 2020] A survey in **Ethiopia** including 1,300 children aged 10 to 23 months reported 798 (61%) had received the measles vaccine. Women who could not read and write were five times more likely to have an incompletely immunised child than those educated with a diploma, degree, and above (Adjusted Odds Ratio = 5.1, 95% CI 2.3 - 11.1). [Wale TA 2020]

Retrospective data from **US** states on local immunisation programs among kindergarten children (age 5-6 years) reported 95% had two doses of measles, mumps, and rubella vaccine in the 2019/20 school year. COVID disruptions were expected to reduce vaccination coverage in the 2020/21 school year. [Seither R 2021] Data from the Michigan Care Improvement Registry in the US reported that disruptions impacted coverage. For 16-month-olds, measles vaccination coverage decreased from 76.1% in May 2019 to 70.9% in May 2020. [Bramer CA 2020] Data from the **US** CDC reported significant reductions in measles vaccines over 15 weeks. The greatest reduction in orders was reported in the week beginning 13 April, with a cumulative decrease of approximately 400,000 measles doses compared to 2019. Smaller declines were seen in children aged ≤24 months than in older children. [Santoli JM 2020]

A review of routine immunisation data from 15 **African** countries showed that thirteen saw drops in the monthly average vaccine doses provided. Nine countries had lower monthly first dose measles vaccinations in the second quarter of 2020 compared to the first quarter. Those countries with more inadequate coverage before COVID-19 reported more significant drops in the number vaccinated after the pandemic was declared. [Masresha BG 2020] A retrospective study in a hospital vaccination unit in Senegal comparing measles vaccinations, amongst others, reported significant reductions in coverage. [Sow 2020] On 15 May 2020, as part of the pulse survey, the WHO reported that measles campaigns were suspended in 27 countries. [**WHO pulse survey 15 May report 2020]**

Electronic health records of children <1-year-old in **England** reported measles-mumps-rubella vaccination was 20% lower in the first three weeks of restrictions than the same period in 2019, before improving in mid-April. [McDonald 2020] The Nuffield Trust reported the number of vaccinations decreased in the week beginning 23 March 2020 (week 13) when lockdown measures began. [Nuffield Trust 2021] Vaccination numbers then increased from week 16 and remained relatively stable at pre-pandemic levels. Public Health **England** [Public Health England 2021] reported that vaccine coverage for MMR1 between September and December 2020 was lower than 2019 estimates and that coverage was already below the WHO target of 95% in 2019. [5] In the **Netherlands**, the first MMR vaccination from March to September 2020 dropped by between 6 and 14% compared with the previous year. After catch-up vaccinations, a difference in the first MMR vaccination of −1% to −2% remained. [Middeldorp M 2021]

There were two modelling studies. Abbas K et al. 2020 modelled a high-impact scenario and a low-impact scenario in **Africa** to approximate child deaths relating to measles that immunisation coverage reductions could cause during COVID-19 outbreaks. Carias C et al. 2021 modelled projected **US** measles vaccination coverage for one-year-olds in 2020 for different durations of stay-at-home orders.

### Polio

A Review of the Global Polio Eradication Initiative (GPEI) reported a halt to polio vaccination until the second half of 2020. GPEI comprises six organisations: the WHO, the US CDC, UNICEF, Rotary International, the Bill & Melinda Gates Foundation, and the Global Alliance for Vaccines and Immunisations. Data from **Afghanistan** reported 21 confirmed polio cases in 2018, 29 in 2019, and, in 2020, 34 confirmed cases were reported as of August 1, 2020. [Ahmadi 2020]

We reviewed the latest **Afghanistan** data from GPEI (https://polioeradication.org/). As of 16 June, one case of wild poliovirus type 1 (WPV1) has been reported for 2021 and a total of 56 cases in 2020. ^**[6]**^ The latest status for Afghanistan suggests that the country remains affected by WPV1 and circulating vaccine-derived poliovirus type 2 (cVDPV2). In March 2021, 6.6 million children were vaccinated against polio during the National Immunisation Days. ^**[7]**^ There were forty cases of circulating cVDPV2 reported in 2021 and 308 cases in 2020.

An overview of the impact of COVID-19 on polio vaccination in **Pakistan** reported that vaccination campaigns in Pakistan were suspended in April 2020. Due to COVID-19-related disruptions to services, 40 million children missed polio vaccinations. [Din M 2020]. In Sindh, **Pakistan**, one in every two children in the province missed their routine vaccinations during the lockdown period. [Chandir S 2020]

We similarly reviewed the latest data on the GPEI from **Pakistan**. In 2020, through September, 73 cases of WPV1 were reported in Pakistan and 62 cases of cVDPV2. Cases in Pakistan dropped to eight in 2017, 12 in 2018, and then increased to 147 in 2019. [Chandir S 2020] The latest data from Pakistan as of June 16 reports one case of WPV1 in 2021 and 84 cases in 2020. The number of cVDPV2 cases so far in 2021 is eight; in 2020, there were 135 cases. In March 2021, a national Immunisation campaign ran from 29 March to 2 April 2021, with 40.1 million children vaccinated. ^**[8]**^

Data from the **Rwanda** Health Management Information System from March and April 2019 compared to 2020 showed significant reductions in polio 1 and 2 vaccinations. [Wanyana 2021] In a hospital centre in **Senegal**, polio vaccination was reduced from March to August 2020. [Sow A 2020] Data from **Sierra Leone** on five common vaccinated diseases from Mar 1, 2020, to Apr 26, 2020, compared with 2019, reported decreases in vaccination ranging from 50 to 85% depending on the individual vaccine analysed, including the OPV1 vaccine. [Buonsenso D 2021]

In April 2020, The WHO reported that **Niger** had an outbreak of vaccine-derived poliovirus that affected two children—having suspended the vaccination campaign due to the pandemic. Niger’s last wild polio case was in 2012 ^**[9]**^. Niger joins 15 countries experiencing vaccine-derived poliovirus outbreaks in Africa. No wild poliovirus has been detected in **Africa** since 2016. Niger joins the list of countries experiencing vaccine-derived poliovirus outbreaks in Africa. The other countries are Angola, Benin, Burkina Faso, Cameroon, Central African Republic, Chad, Côte d’Ivoire, the Democratic Republic of the Congo, Ethiopia, Ghana, Mali, Nigeria, Togo and Zambia.

## Discussion

The evidence in this review comes from a wide range of conditions and countries that highlight significant disruptions to childhood vaccine services. We found reductions in diphtheria tetanus pertussis, BCG, measles and polio vaccine uptake, as well as a range of other vaccines regularly given to children. Reductions in many settings were reversed once restrictions were lifted. However, not all settings reported a return to complete normality. Smaller declines were seen in younger children than older children. In addition, children born to women who could not read and write were more likely to be incompletely immunised. Barriers to access and maintaining public transport infrastructure impacted uptake of childhood vaccinations as well.

The true impact of COVID-19 disruptions on childhood vaccination services is yet to be determined. Two years after reductions in measles vaccination during the Ebola outbreak in Guinea, the incidence of measles increased from 2.7 per million in 2015 to 11.5 per million in 2016 and 52.5 per million in 2017, of which 65% of cases were confirmed in those aged < 5 years. ^**[10]**^ Worldwide measles deaths climbed by 50% from 2016 to 2019, with over 200,000 lives lost in 2019. ^**[11]**^

Low levels of polio vaccination and a lack of immunity can leave countries at risk of polio returning. Indonesia, Mozambique, Myanmar, Papua New Guinea, and the Philippines are currently considered key-at-risk countries by GPEI, Afghanistan, and Pakistan affected by ongoing endemic WPV1 and cVDPV2. ^[11]^ There is a concerted effort underway to eradicate polio. The GPEI reports fewer cases in 2021 compared to the same period last year. This is promising, despite the lockdowns; however, the potential for underreporting exists due to the pandemic. The effects of Polio can be devastating: in 1996, wild poliovirus paralysed more than 75,000 children in Africa. ^[12]^ Therefore, maintaining sustained vaccination levels and disease surveillance remains a priority for at-risk countries. Furthermore, several barriers were reported to vaccination, including a lack of clarity around whether vaccination services were operating, as usual, difficulties in organising vaccination appointments, and fears around contracting COVID-19. [Bell S 2020]

## Limitations

We did not formally assess the quality of the included studies. However, clinical audit and service evaluation remain valuable for analysing and targeting improvements in healthcare. Publication bias will favour studies that highlight disruptions and account for excess reporting of decreases in vaccination levels. However, alongside those studies, reviews by international bodies highlight consistent drops in vaccination uptakes during the restrictive phases of the pandemic. Studies that report no reductions in vaccination are essential; they highlight strategies that can overcome disruptions during restrictions and – crucially - after they have been lifted. Reports of vaccine coverage may not be 100% accurate, particularly during periods of upheaval such as in a pandemic.

## Conclusions

COVID-19 pandemic measures caused significant disruption to childhood vaccination services and uptake. In future pandemics, and for the remainder of the current one, policymakers must ensure access to vaccination services and provide catch-up programs to maintain high levels of immunisation, especially in those most vulnerable to childhood diseases in order to avoid further inequalities.

## Data Availability

All data included in the review is provided in the tables and text.

## Acknowledgements

The authors would like to thank Martin Kulldorff and AJ Kay for comments on the draft.

## Ethics Committee Approval

No approval was necessary.

## Data Availability

All data included in the review is provided in the tables and text.

## Appendices

**References to Included Studies, Tables 1 & 2:**

**Primary Studies on Childhood Vaccination Uptake During the COVID-19 Pandemic (TABLE 1)**

**Reports by National and International Agencies on Childhood Vaccination Uptake During the COVID-19 Pandemic (TABLE 2)**

